# COVID-19 diagnosis prediction in emergency care patients: a machine learning approach

**DOI:** 10.1101/2020.04.04.20052092

**Authors:** André Filipe de Moraes Batista, João Luiz Miraglia, Thiago Henrique Rizzi Donato, Alexandre Dias Porto Chiavegatto Filho

## Abstract

The coronavirus disease (COVID-19) pandemic has increased the necessity of immediate clinical decisions and effective usage of healthcare resources. Currently, the most validated diagnosis test for COVID-19 (RT-PCR) is in shortage in most developing countries, which may increase infection rates and delay important preventive measures. The objective of this study was to predict the risk of positive COVID-19 diagnosis with machine learning, using as predictors only results from emergency care admission exams. We collected data from 235 adult patients from the Hospital Israelita Albert Einstein in São Paulo, Brazil, from 17 to 30 of March, 2020, of which 102 (43%) received a positive diagnosis of COVID-19 from RT-PCR tests. Five machine learning algorithms (neural networks, random forests, gradient boosting trees, logistic regression and support vector machines) were trained on a random sample of 70% of the patients, and performance was tested on new unseen data (30%). The best predictive performance was obtained by the support vector machines algorithm (AUC: 0.85; Sensitivity: 0.68; Specificity: 0.85; Brier Score: 0.16). The three most important variables for the predictive performance of the algorithm were the number of lymphocytes, leukocytes and eosinophils, respectively. In conclusion, we found that targeted decisions for receiving COVID-19 tests using only routinely-collected data is a promising new area with the use of machine learning algorithms.

## Introduction

During the last month, coronavirus disease 2019 (COVID-19) has spread rapidly from China to European countries, and is now experiencing exponential growth in the United States and Canada.^1^ Recently, there has also been a sharp increase in cases in developing countries, but due to a limited number of COVID-19 tests available for these countries, the real extent of the spread is still unknown, which may lead to dire consequences in terms of adequate clinical care and the epidemiological knowledge needed to guide public containment measures.^2,3^

The most common symptoms of COVID-19, such as fever and cough, are similar to a range of other infectious diseases, making prompt diagnosis a challenge for health professionals.^4^ Results from the transcription polymerase chain reaction test (RT-PCR), currently the most reliable diagnostic test, are frequently taking more than a week to become available according to reports in the Brazilian media, while in the meantime there is a need for immediate decisions about clinical care and preventive measures. On the other hand, the recent increase in the usage of new rapid diagnostic tests, which are prone to some accuracy issues, may increase the risk of inefficient allocations of health resources.

This study aims to improve decisions regarding COVID-19 test priorities in developing countries by predicting the risk of a positive diagnosis, using only routinely-collected data from emergency care admission exams. We analyzed the results of COVID-19 tests performed at Hospital Israelita Albert Einstein (HIAE) in São Paulo, Brazil, one of the main test providers in the country during the first weeks of local outbreak. This study was conducted under a task-force established to respond to the COVID-19 emergency within the PROADI-SUS program.

## Methods

According to current guidelines from the Brazilian Ministry of Health, COVID-19 tests should only be applied to patients with severe acute respiratory syndrome.^5^ Between 17 and 30 of March 2020, there were 256 results for COVID-19 transcription polymerase chain reaction (RT-PCR) tests performed in patients admitted to the emergency department of HIAE with a corresponding blood count test. A total of 235 adults had full results for total blood count and polymerase chain reaction (PCR) in the 24 hours around the RT-PCR test and were included in the analysis. From these, 102 (43%) patients tested positive for COVID-19.

A total of 15 variables were used for training the algorithms: age, gender, hemoglobin, platelets, red blood cells, mean corpuscular hemoglobin concentration (MCHC), mean corpuscular hemoglobin (MCH), red cell distribution width (RDW), mean corpuscular volume (MCV), leukocytes, lymphocytes, monocytes, basophils, eosinophils and c-reactive protein (CRP).

We tested the predictive performance for positive diagnosis of COVID-19 of five popular machine learning algorithms (neural networks, gradient boosted trees, random forests, logistic regression and support vector machines) with 10-fold cross validation for hyperparameter tuning with hyperopt (Bayesian optimization) search. All variables except gender were numerical and were therefore normalized to avoid oversized effects due to difference in scales. The sample was randomly divided using a 70-30 split, where 70% of the patients were used to train the machine learning algorithms, and the other 30% were used to test the performance of the models on new unseen data. All the results presented here are from the test set.

We measured predictive performance by calculating the area under the ROC curve (AUC), sensitivity, specificity, F1-score, Brier score, positive predictive value (PPV) and negative predictive value (NPV). All analyses were performed in Python using the scikit-learn library. The study was performed in accordance with the guidelines of Transparent Reporting of a Multivariable Prediction Model for Individual Prognosis or Diagnosis (TRIPOD) whenever applicable.^6^ The study was approved by the Hospital Israelita Albert Einstein IRB (project number 4110-20) and by the National Commission for Ethics in Research (CONEP) of the National Health Council (CNS) from the Ministry of Health (CAAE: 30414720.0.0000.0071).

## Results

Table 1 presents the descriptive results for the features included in the models, for all patients and separated according to COVID-19 diagnosis. The full sample was well balanced between males and females (51.1% and 48.9%, respectively), with a mean age of 49 years old. Within the COVID-19 positive group there were more men (65.7%), and lower mean values for leukocytes, lymphocytes, monocytes, basophil and eosinophils.

**Table 1.**
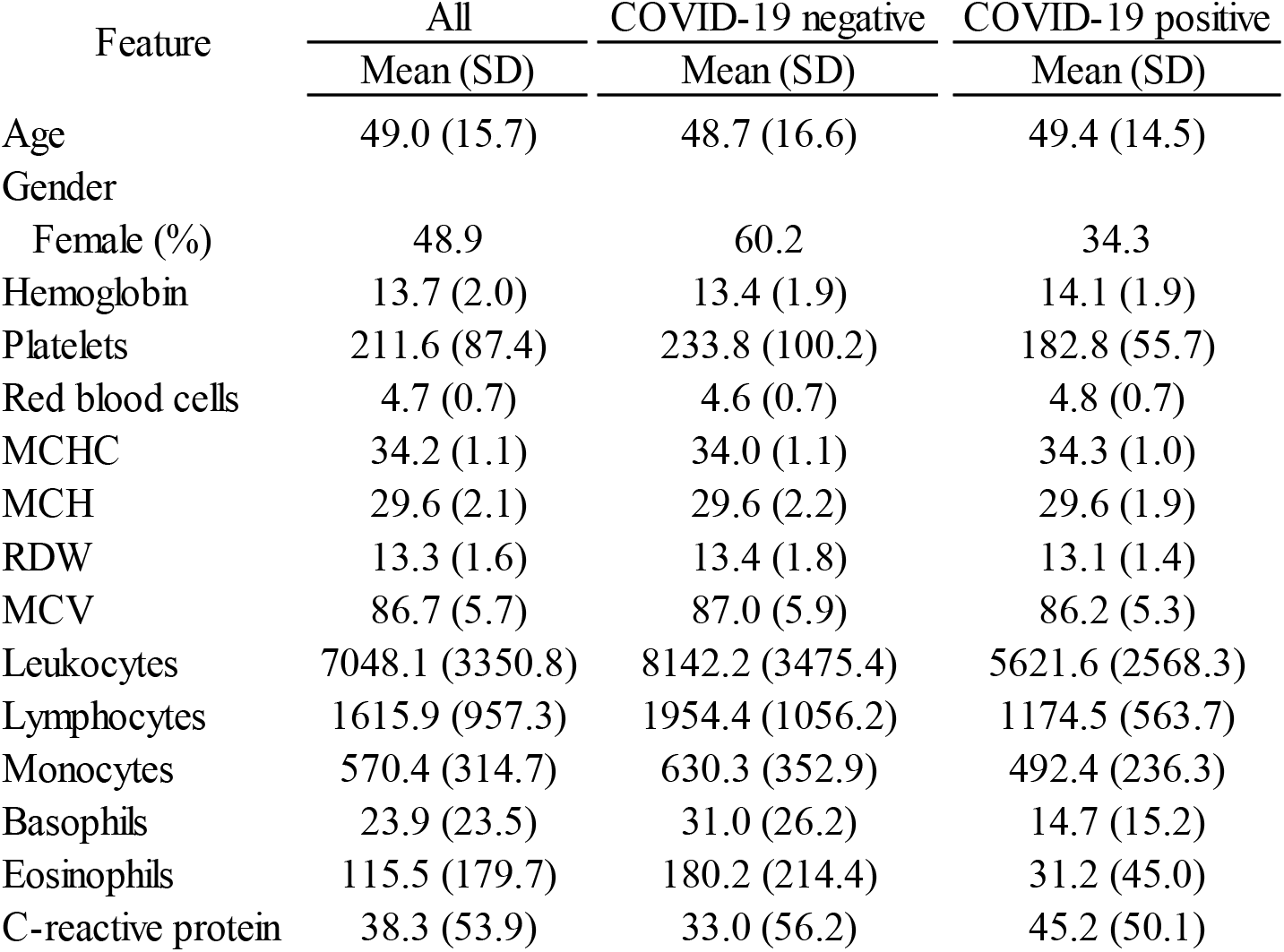
Descriptive results for the features included in the models, for the complete sample and separated by COVID-19 diagnosis.

| Feature | All | COVID-19 negative | COVID-19 positive |
| --- | --- | --- | --- |
|  | Mean (SD) | Mean (SD) | Mean (SD) |
| Age | 49.0 (15.7) | 48.7 (16.6) | 49.4 (14.5) |
| Gender |  |  |  |
| Female (%) | 48.9 | 60.2 | 34.3 |
| Hemoglobin | 13.7 (2.0) | 13.4 (1.9) | 14.1 (1.9) |
| Platelets | 211.6 (87.4) | 233.8 (100.2) | 182.8 (55.7) |
| Red blood cells | 4.7 (0.7) | 4.6 (0.7) | 4.8 (0.7) |
| MCHC | 34.2 (1.1) | 34.0 (1.1) | 34.3 (1.0) |
| MCH | 29.6 (2.1) | 29.6 (2.2) | 29.6 (1.9) |
| RDW | 13.3 (1.6) | 13.4 (1.8) | 13.1 (1.4) |
| MCV | 86.7 (5.7) | 87.0 (5.9) | 86.2 (5.3) |
| Leukocytes | 7048.1 (3350.8) | 8142.2 (3475.4) | 5621.6 (2568.3) |
| Lymphocytes | 1615.9 (957.3) | 1954.4 (1056.2) | 1174.5 (563.7) |
| Monocytes | 570.4 (314.7) | 630.3 (352.9) | 492.4 (236.3) |
| Basophils | 23.9 (23.5) | 31.0 (26.2) | 14.7 (15.2) |
| Eosinophils | 115.5 (179.7) | 180.2 (214.4) | 31.2 (45.0) |
| C-reactive protein | 38.3 (53.9) | 33.0 (56.2) | 45.2 (50.1) |

The two algorithms with best overall performance were random forests and support vector machines with an AUC of 0.85 for the test set. Figure 1 presents the AUC of the five algorithms, which shows that the performance was similar between the algorithms with some overlap across the different discriminative thresholds.

**Figure 1:**
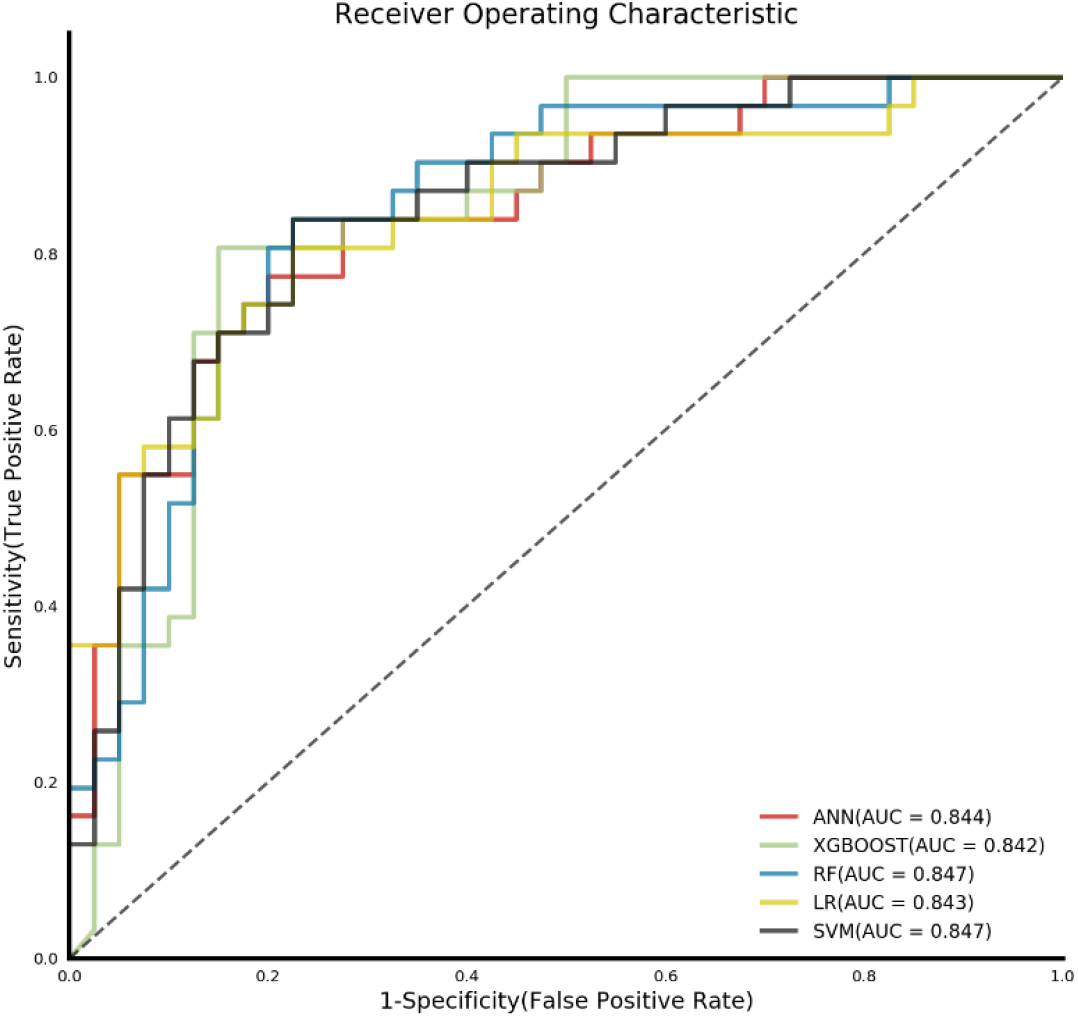
Receiver Operating Characteristic (ROC) curve for the test set of each of the five machine learning algorithms.

The two algorithms with the best predictive performance (support vector machines and random forests) had the same discrimination results (sensitivity of 0.677 and specificity of 0.850), but the support vector machines algorithm had a slightly better calibration, with a Brier score of

0.160 (Table 2). Every one of the five algorithm had a positive predictive value of at least 0.74 and a negative predictive value of at least 0.77.

**Table 2:** Performance metrics for each machine learning algorithm for the test set.

| Algorithm | AUC | Sensitivity | Specificity | F1-score | Brier score | PPV | NPV |
| --- | --- | --- | --- | --- | --- | --- | --- |
| Supp. Vec. Machines | 0.847 | 0.677 | 0.850 | 0.724 | 0.160 | 0.778 | 0.773 |
| Random Forests | 0.847 | 0.677 | 0.850 | 0.724 | 0.161 | 0.778 | 0.773 |
| Neural Networks | 0.844 | 0.742 | 0.800 | 0.742 | 0.187 | 0.742 | 0.800 |
| Logistic Regression | 0.843 | 0.742 | 0.825 | 0.754 | 0.161 | 0.767 | 0.805 |
| Grad. Boost. Trees | 0.842 | 0.806 | 0.800 | 0.781 | 0.171 | 0.758 | 0.843 |

Calibration details for the support vector machines algorithm are presented in Figure 2, which shows that despite the small sample size, the algorithm is well calibrated throughout the probability distribution, i.e. among patients with a high predicted risk according to the algorithm, a high percentage of them are COVID-19 positive, and vice-versa. The five most important variables for the predictive performance of the algorithm according to the Mean Decrease Accuracy (MDA) were lymphocytes, leukocytes, eosinophils, basophils and hemoglobin, respectively.

**Figure 2:**
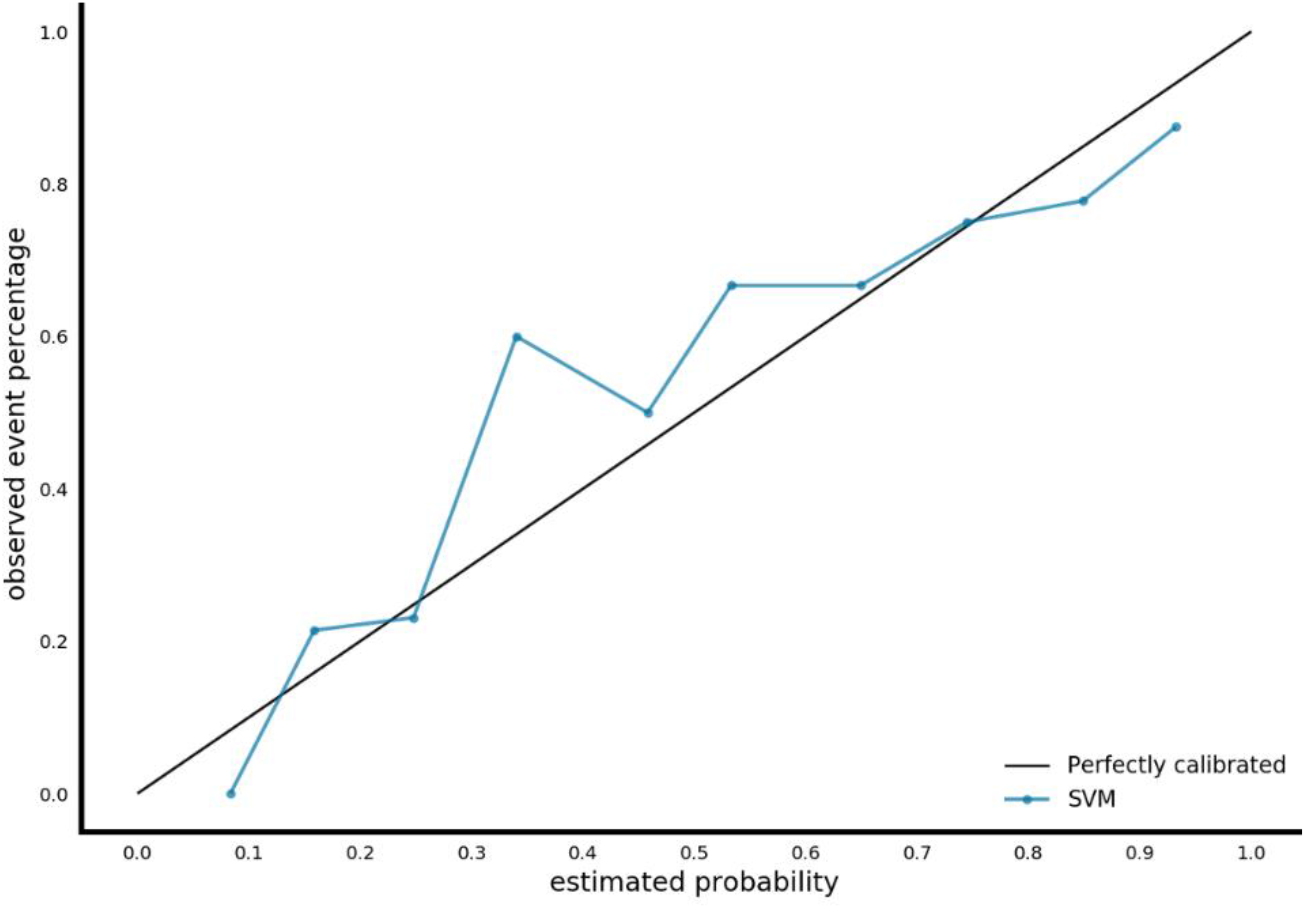
Calibration results for the support vector machine algorithm on the test set.

We also tested the performance of the algorithms using 10-fold cross validation repeated 10 times for each algorithm, instead of using the 70-30 split. This strategy is frequently used in machine learning in health studies especially in cases of relatively small samples such as ours, but it simulates less clearly the results in clinical applications. The predictive performance of the algorithms was better in comparison with the previous strategy, with an AUC of 0.87, Sensitivity of 0.755 and Specificity of 0.829 for the best-performing algorithm, which was again the support vector machines (Annex 1).

## Discussion

We found that by using only standard exams performed upon emergency care admission, machine learning algorithms were able to predict with good performance the risk of each patient having a positive result for COVID-19. As of April 5, 2020, there have been a total of 11,130 confirmed cases of COVID-19 in Brazil. Due to an overall shortage of tests, the current recommendation from Brazilian Ministry of Health is that tests should only be performed for critically-ill patients, contrary to the guidance of the World Health Organization which encourages large-scale tests of the population.^7^ There are now also confirmed cases in most African countries and India, where the potential of rapid spread will require harsher cost-effective decisions on which patients to test for COVID-10.

The five machine learning algorithms had similar predictive performances, with AUCs higher than 0.84. The calibration results for the best performing algorithm (support vector machines) was consistent throughout the entire probability distribution. For example, among the patients with a predicted probability between 80 and 100% of having COVID-19, 82% of them actually had it; while for those with a predicted probability of 0 to 20%, 12% had it. This means future use of predicted probabilities is also promising, not only the binary decision of having COVID-19 or not, which improves the strength of potential clinical decisions derived from the results.

During the last few weeks, the first studies were published that apply machine learning to predict different COVID-19 outcomes.^8^ So far only one study analyzed diagnosis of COVID- 19 with routinely-collected data, in this case to predict cases using as controls patients with viral pneumonia by applying logistic regression.^9^ Another study of 53 patients from two hospitals in Wenzhou, China, analyzed the accuracy of five machine learning algorithms to predict Acute Respiratory Distress Syndrome (ARDS) in patients with COVID-19.^10^ Two other studies applied machine learning algorithms to predict mortality in patients with COVID-19, using patient data from Kaggle and China.^11,12^

We propose that machine learning algorithms can be used to allocate priorities in receiving the RT-PCR tests in the case of a shortage, and also to help with critical care decisions while the RT-PCR results are being processed (which have been frequently taking more than a week in most places of Brazil). A promising area for future research will also be to analyze the combined performance of the new rapid tests and the machine learning algorithms.

Further studies need to be performed in other locations to confirm these results. We analyzed a relatively small sample from the Hospital Israelita Albert Einstein, which was a first-responder for COVID-19 cases in Brazil, but that has a large proportion of high-income patients and is frequently considered among the top hospitals in Latin America. Different stages of the disease may also influence the predictive performance of the models. We tried to decrease this effect by analyzing only patients in emergency care, but time interval from the first symptoms may also be a factor.

In conclusion, we found evidence to suggest that targeted decisions for receiving COVID- 19 tests in areas with a shortage of supply is possible with the use of machine learning algorithms. For future clinical applications, locally training the algorithms in each specific healthcare service will be crucial to improve predictive performance.

## Data Availability

Due to the nature of this research, participants of this study did not agree to share publicly their individual data, so supporting data is not available.

## Acknowledgements

We thank the support of the Big Data Analytics team of the Hospital Israelita Albert Einstein, especially Eduardo Reis, Ellison Fernando, Rodrigo Carvalho and Johnatha Franca, that helped with the preparation and availability of the database.

---

Annex 1 - Performance metrics for each machine learning algorithm for the 10-fold cross- validation.

| Algorithm | AUC | Sensitivity | Specificity | F1-score | Brier score | PPV | NPV |
| --- | --- | --- | --- | --- | --- | --- | --- |
| SVM | 0.866 | 0.755 | 0.829 | 0.761 | 0.145 | 0.784 | 0.822 |
| Grad. Boost. Trees | 0.862 | 0.878 | 0.700 | 0.774 | 0.167 | 0.701 | 0.889 |
| Random Forests | 0.861 | 0.753 | 0.846 | 0.767 | 0.153 | 0.798 | 0.825 |
| Logistic Regression | 0.853 | 0.780 | 0.774 | 0.750 | 0.153 | 0.737 | 0.829 |
| Neural Networks | 0.845 | 0.747 | 0.796 | 0.738 | 0.169 | 0.749 | 0.808 |

